# Chromatic fusion: generative multimodal neuroimaging data fusion provides multi-informed insights into schizophrenia

**DOI:** 10.1101/2023.05.18.23290184

**Authors:** Eloy P.T. Geenjaar, Noah L. Lewis, Alex Fedorov, Lei Wu, Judith M. Ford, Adrian Preda, Sergey M. Plis, Vince D. Calhoun

**Author notes:** Correspondence Eloy Geenjaar. **Funding information**.

## Abstract

This work proposes a novel generative multimodal approach to jointly analyze multimodal data while linking the multimodal information to colors. By linking colors to private and shared information from modalities, we introduce chromatic fusion, a framework that allows for intuitively interpreting multimodal data. We test our framework on structural, functional, and diffusion modality pairs. In this framework, we use a multimodal variational autoencoder to learn separate latent subspaces; a private space for each modality, and a shared space between both modalities. These subspaces are then used to cluster subjects, and colored based on their distance from the variational prior, to obtain meta-chromatic patterns (MCPs). Each subspace corresponds to a different color, red is the private space of the first modality, green is the shared space, and blue is the private space of the second modality. We further analyze the most schizophrenia-enriched MCPs for each modality pair and find that distinct schizophrenia subgroups are captured by schizophrenia-enriched MCPs for different modality pairs, emphasizing schizophrenia’s heterogeneity. For the FA-sFNC, sMRI-ICA, and sMRI-ICA MCPs, we generally find decreased fractional corpus callosum anisotropy and decreased spatial ICA map and voxel-based morphometry strength in the superior frontal lobe for schizophrenia patients. To additionally highlight the importance of the shared space between modalities, we perform a robustness analysis of the latent dimensions in the shared space across folds. These robust latent dimensions are subsequently correlated with schizophrenia to reveal that for each modality pair, multiple shared latent dimensions strongly correlate with schizophrenia. In particular, for FA-sFNC and sMRI-sFNC shared latent dimensions, we respectively observe a reduction in the modularity of the functional connectivity and a decrease in visual-sensorimotor connectivity for schizophrenia patients. The reduction in modularity couples with increased fractional anisotropy in the left part of the cerebellum dorsally. The reduction in the visual-sensorimotor connectivity couples with a reduction in the voxel-based morphometry generally but increased dorsal cerebellum voxel-based morphometry. Since the modalities are trained jointly, we can also use the shared space to try and reconstruct one modality from the other. We show that cross-reconstruction is possible with our network and is generally much better than depending on the variational prior. In sum, we introduce a powerful new multimodal neuroimaging framework designed to provide a rich and intuitive understanding of the data that we hope challenges the reader to think differently about how modalities interact.

## 1 INTRODUCTION

The acquisition of neuroimaging data often consists of various modalities, such as structural, functional, and diffusion magnetic resonance imaging data. Access to a wide range of complementary modalities is necessary to deeply understand the relationship between (functional/structural) brain patterns and demographic/neuropsychiatric variables [1]. For example, unimodal studies may show that brain activity and structure are both linked to the same neuropsychiatric variable. This, however, does not allow us to draw the conclusion that both modalities (linearly or nonlinearly) covary together. In fact, their specific covariation may have an even stronger link to the neuropsychiatric variable under study. Understanding the covariation pattern of two modalities and how that covariation relates to neuropsychiatric variables also allows us to dig deeper into the latent mechanism that underlies both the divergence in behavioral and neuroimaging measures. When covariation in divergent brain activity and structure are linked to the same brain region, the effects may be localized. However, when covariation indicates different brain regions for each modality, there may be a more complex mechanism underlying their effect on the neuropsychiatric variable. Importantly, jointly estimating the multimodal relationships can allow the modalities to inform one another, yielding information that would be individual to a unimodal analysis. A rich understanding of these patterns will lead to complementary quantitative visualizations or measures that can have the potential to help clinicians make decisions about diagnosis and treatment plans. Further, access to quantitative visualizations will ultimately be important to help practitioners facilitate the delivery of personalized medicine [2].

Multimodal neuroimaging is thus a critical aspect of studying the brain and aims to incorporate each modality as a piece of the puzzle to obtain a complete picture of the brain. Just as with pieces of a puzzle, modalities have shared information; the shape of a puzzle piece determines how it fits with certain other puzzle pieces and exclusive or private information; is the picture printed on the puzzle piece. To separate out the shared and private information from multimodal datasets, recent work in deep learning has started to develop neural networks that extract these two types of information [3, 4]. Specifically, our multimodal neuroimaging model leverages and extends the disentangled multimodal variational autoencoder (DMVAE) [4] as a building block. The goal of our work is to learn lower-dimensional representations of multimodal datasets in such a way that we can understand how the interactions among the modalities in each dataset relate to schizophrenia. We chose to use an approach that models two modalities as having private and shared information because it induces a structure in our lower-dimensional representations that both constrains the solution space of our algorithm and is interpretable. Furthermore, lower-dimensional representations allow us to summarize each modality in a few important factors of variation, and the use of an autoencoder ensures that we have access to a decoder that can map these important factors of variation back into the original space of the modality to aid in the interpretation of the results. We expect that explicitly modelling the shared information between modalities with a non-linear method can lead to interesting patterns between those shared factors and schizophrenia. Specifically, when interpolating along certain shared schizophrenia-enriched factors, we expect non-linear covariations to relate to schizophrenia.

Moving towards multi-dimensional (continuous) measures to understand psychiatric disorders is important because binary representations can be misleading. For example, schizophrenia often co-occurs with other mental disorders [5]. Additionally, there is significant intra-diagnostic heterogeneity for schizophrenia, and the lines between other severe mental disorders remain blurred. Thus, representing features of an individual’s brain on a continuous multi-dimensional spectrum is important to understand individual brain differences, and predict risk [6]. Integrating multiple modalities and viewing subjects on a spectrum is consistent with the trans-diagnostic NIMH research domain criteria (RDoC) initiative [7].

In this work, we propose a new framework to analyze spectrum psychiatric disorders through a powerful multimodal representation learning framework. Specifically, we disentangle private and shared information from a pair of modalities and characterize schizophrenia patients from the perspective of these distinct forms of information. Namely, given their variational inference framework, variational autoencoders enforce a certain prior during training. In our case, we use a zero-mean normal distribution, which allows us to naturally interpret encoded modalities further from this prior than others to be more irregular in the dataset. Based on the three distinct types of information; the private information from the first and second modalities, and their shared information, we can assign a color to each cluster based on how irregular each of these types of information is for that specific subject. Considering there are three axes along which the color can change, we use a red-green-blue (RGB) color model to assign a color to each cluster, where red and blue represent private information from the first and second modalities, respectively, and green represents their shared information. For example, a subject whose two modalities contradict each other in terms of encoded information will be irregular according to their shared information and have a high value for green. We call this framework of assigning a color to clusters based on the information encoded in their modalities: chromatic fusion.

In our analyses, we use all potential pairs of four modalities; spatial ICA maps, static functional connectivity (sFNC), fractional anisotropy maps (FA), and voxel-based morphometry (sMRI). Although sFNC and spatial ICA maps are derived from the same modality, namely functional magnetic resonance imaging (fMRI), we refer to them as different modalities in this manuscript for brevity. Our goal is to both introduce a new way of thinking about combining modalities, and visualizing how those combinations of modalities uniquely highlight subgroups and thus the heterogeneity of psychiatric spectrum disorders, such as schizophrenia. Specifically, for each modality pair, we look at naturally arising significantly schizophrenia-enriched clusters; clusters with a large number of schizophrenia subjects, whether those clusters capture the same subjects across modality pairs, and what patterns in the data these clusters represent. Hence, we hypothesize that different modality pairs lead to distinct patterns related to schizophrenia. We aim to find these patterns without supervised labels and compare them across modality pairs. In fact, we show that these naturally arising clusters are a function of which modalities are paired. Our results imply that distinct combinations of modalities are able to highlight different schizophrenia subgroups in our dataset. This also indicates how heterogeneity in schizophrenia is a function of the interactions between modalities. Meaning that different subgroups are distinct from the rest of the sample based on which modalities are combined. Additionally, we show that latent dimensions compromising the shared information space are significantly related to schizophrenia. We include analyses on the shared latent dimensions, how robust these shared latent dimensions are, and whether they are correlated with schizophrenia. Generally, we find robust and correlated shared dimensions for each modality pair, and highlight two to visualize unique and multimodal patterns significantly related to schizophrenia.

### 1.1 Previous work

In multimodal machine learning one of the main challenges is to learn good representations [8]. Good representations are essentially accurate and interpretable summaries of a data sample at hand. An example of an impactful summary of neuroimaging data is independent component analysis (ICA) spatial maps or functional connectivity [9, 10]. Our method addresses this challenge and performs symmetric multimodal fusion. As opposed to asymmetric fusion, where one modality constrains another, we analyze two modalities concurrently in a symmetric fusion model. Furthermore, we look at how the sub-elements of the modalities are fused after training, and summarize (possibly nonlinear) covariations in the data, instead of trying to find mechanistic relationships between modalities. There are also important distinctions within the field of multimodal data fusion itself [1]. Mainly, there are model-driven and data-driven approaches. Model-driven approaches have the advantage of specifying apriori hypotheses that can be tested, and that those hypotheses can be directional. However, if any important hypotheses are missed, these methods can make incorrect inferences. Data-driven approaches on the other hand do not require the specification of a priori hypotheses. Our model falls under data-driven multimodal fusion approaches. Lastly, there is a distinction between blind and semi-blind data-driven fusion approaches [1]. Semi-blind approaches, in contrast to blind approaches, use some a priori knowledge to constrain the solution space of the model, such as regularization. As discussed in the previous paragraph, by encouraging our model to find private and shared information for two modalities, we constrain the solution space of the model. Thus, our approach falls under the umbrella of unsupervised semi-blind multimodal fusion approaches. Examples of other semi-blind multimodal fusion approaches are joint ICA [11], which assumes a shared loading parameter, multiset canonical correlation analysis with reference + joint ICA (mCCAR+jICA) [12], which allows a behavioral reference to constrain the solution, independent vector analysis (IVA) [13, 14, 15], which assumes an independent/dependence structure to extract linked sources, and parallel ICA, which optimizes jointly for independence within a modality, and linear covariation between a subset of the sources. These multimodal fusion approaches are complex and thus indicate there is a potential benefit in leveraging the flexibility of deep learning models in the context of multimodal fusion. Further, we want to move beyond the linear mixing assumption and explicitly find private and shared factors among pairs of modalities.

Two recent studies [16, 17], similar to our approach, model multimodal data in terms of its private and shared factors. The authors in the former perform rigorous experiments to find shared and common factors using advanced coupled matrix tensor factorization (ACMTF) to predict whether a task is being performed in time windows of simultaneous electroencephalogram-functional magnetic resonance imaging (EEG-fMRI) data. The work extends previous tensor decomposition approaches that have been successful at finding shared features between modalities and fusing them [18, 19]. The latter of the two previously mentioned studies uses an autoencoder-based model to disentangle the private and shared factors but trains it using a supervised signal to predict infant age. These methods differ from our work both in their goal and approaches. For example, although [17] uses a variational autoencoder as well, they train the model adversarially with supervised signals. Our work is completely unsupervised yet also results in common interpretable patterns in the dataset. In addition, no work has to our knowledge exploited these multimodal relationships to capture subgroups and visualized the continuum of multimodal combinations as in our chromatic/MCP approach.

Other ways of modeling multimodal neuroimaging data have been proposed as well, two recent approaches involve graph neural networks [20, 21] to learn multimodal representations from functional magnetic resonance imaging (fMRI) and diffusion MRI (dMRI) data. Alternatively, previous studies have modeled the interaction between fMRI data and structural MRI (sMRI) data by aligning sMRI components with dynamic functional connectivity [22]. Multimodal learning with sMRI and fMRI data has also been explored using self-supervised learning [23, 24], by minimizing the divergence of representations from the two different modalities. Our work extends these studies by allowing new points to be decoded and visualized within a meta-chromatic space. Our can reveal novel multimodal insights, for example, our results unify co-variations of fractional anisotropy and voxel-based morphometry with increased visual-visual and reduced visual-sensorimotor functional connecitivity, respectively. Furthermore, our approach enables a new way of thinking about modalities in terms of a chromatic framework, which we show can provide additional insights and increase data transparency.

## 2 MATERIALS AND METHOD

### 2.1 Problem setting

Our goals are to 1) build a generative model that allows us to find private and shared latent factors for each modality, and 2) provide a visual representation of the results to facilitate the discovery of additional insights into the relationship between modalities for each subject and how it differs from other subjects across those modalities. To do this, we use the identified factors to estimate colors in the latent space and create a chromatically fused color space to describe the dataset. The type of dataset we study in this work 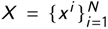 consists of *N* subjects, with *M* modalities per subject 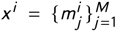 . In this work, we consider the case for *M* = 2. One widespread generative model to tackle this problem is the variational autoencoder (VAE) [25], which maximizes the log-likelihood of reconstructions in the dataset.

The data in this problem is assumed to be sampled from an underlying distribution *p* (z). Throughout this work, we indicate red as private information from the first modality, blue as private information from the second modality, and green as shared information. Colors that are combinations of these colors, e.g. purple is a combination of red and blue, indicate that variables are a combination of the private information of the two modalities. The two modalities are then sampled from the following two conditional distributions, one for each modality 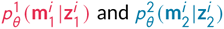 Where 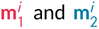 are the two observed modality samples for subject *i* . The marginal distributions for either of these is intractable because of their form; 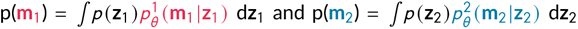 The intractability of these marginal distributions also leads to the intractability of the following true posterior densities: 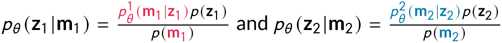. So instead of optimizing over the true posterior densities, the VAE approximates the posterior density for both modalities using an encoder for each 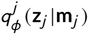. This encoder parameterizes a simpler distribution than the true posterior, in our case an axis-aligned Gaussian distribution. The approximate posterior is variationally optimized to be close to this prior, with zero-mean and diagonal unit covariance. By reparametrizing the conditional distributions, we obtain a variational lower bound on the marginal likelihood of each data point 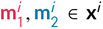. In the case of VAEs this lower bound is called the evidence lower bound (ELBO) and a more in-depth derivation can be found in Kingma and Welling [25]. In our case, we can write the following equation for the ELBO:

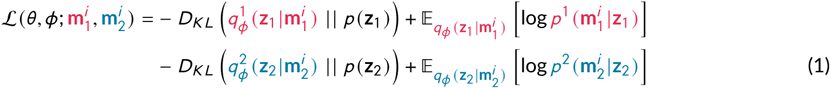

This objective function however does not account for the interactions between modalities. It optimizes the ELBO for the two modalities separately with two separate encoder-decoder models for each modality. To model the shared information between the two modalities, we model those interactions using a disentangled multimodal VAE (DMVAE) [4]. A visual depiction of the DMVAE is shown in Figure 1b. One of the main conceptual ideas behind the DMVAE [4] and other multimodal models that have recently gained traction [3] is the separation of the latent space into a private and shared component for each modality [8].

**FIGURE 1.**
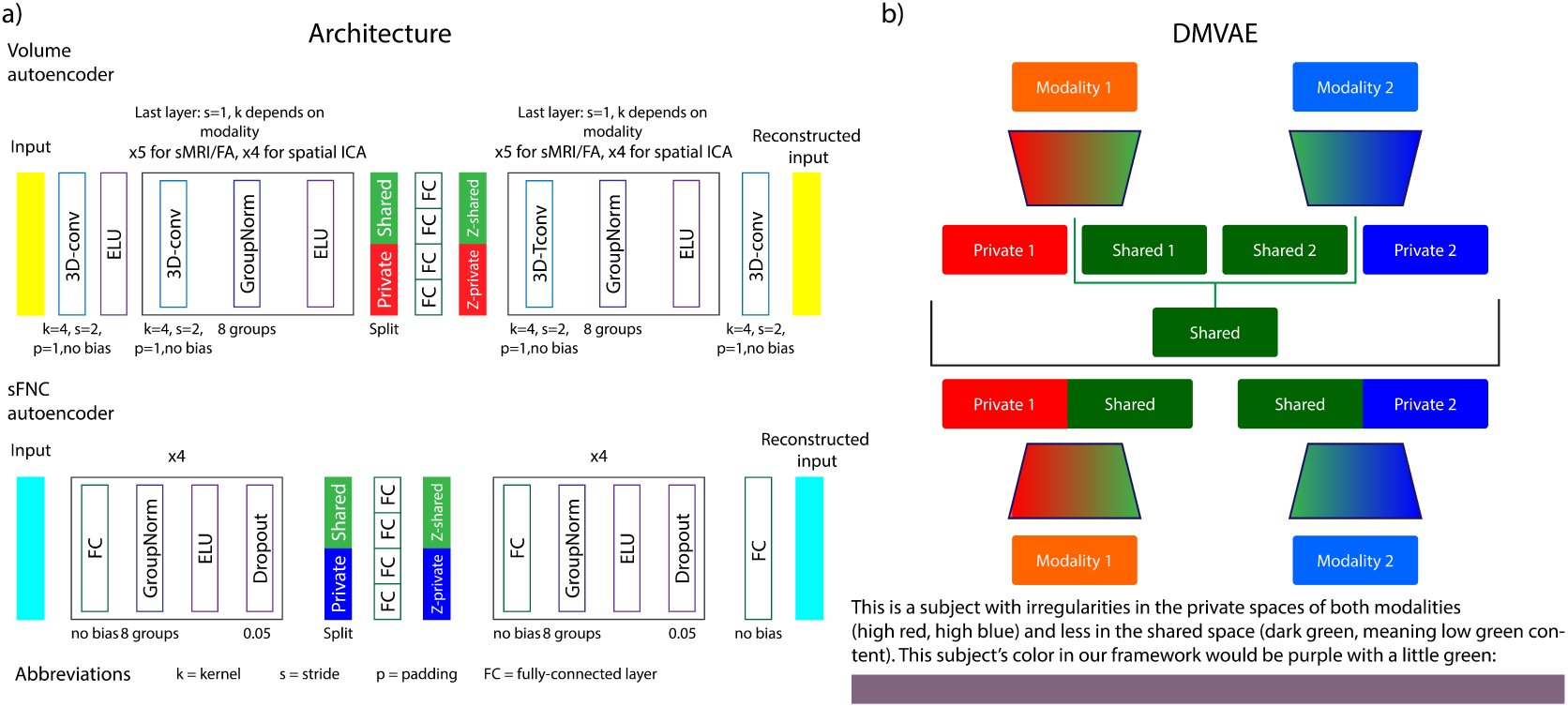
The neural network architecture used for each of the modalities. The left part of this diagram shows the architecture of the encoders and decoders used in this work, and the right part shows the high-level structure of the DMVAE and a visual example of how it extracts the colors of a subject’s modalities into its base colors.

Conceptually, some of the information that is captured by different modalities is mutually exclusive, while other information is shared across modalities. The mutually exclusive information a modality captures is generally referred to as its ’private’ information. For example, T1-weighted structural MRI’s (sMRI) can measure cortical thickness in gray matter as its private information, whereas dMRI can act as an index of axonal organization and coherence in white matter mostly [26]. These modalities, however, share information regarding white matter. We want to ensure we explicitly model modalities this way to disentangle both private and shared information. Throughout the text, we will refer to the private space of subject *i* ’s first modality as 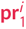, its shared space as sh^*i*^, and the private space of its second modality as 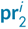 . The encoders will still parameterize Gaussian distributions, but their dimensions will be split into a private and shared space, see Figure 1b. Thus, we obtain two different shared spaces, one for each modality, namely 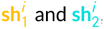, these are colored yellow and cyan in the text because they are a mix between red and green (modality 1 and shared), and blue and green (modality 2 and shared) respectively. To mix these two shared spaces during training, the authors propose to use a product of experts (PoE) [27]. This allows us to obtain a closed form solution for the variance σ_sh^*i*^_ andmean *μ*_sh^*i*^_ of the combined shared distribution: 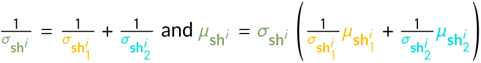. The private and shared dimensions can be expressed in terms of the encoder q (·) as follows: 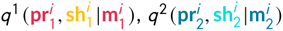 and 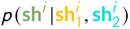. The new objective function then consists of 6 reconstruction and 5 KL-divergence terms. 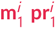

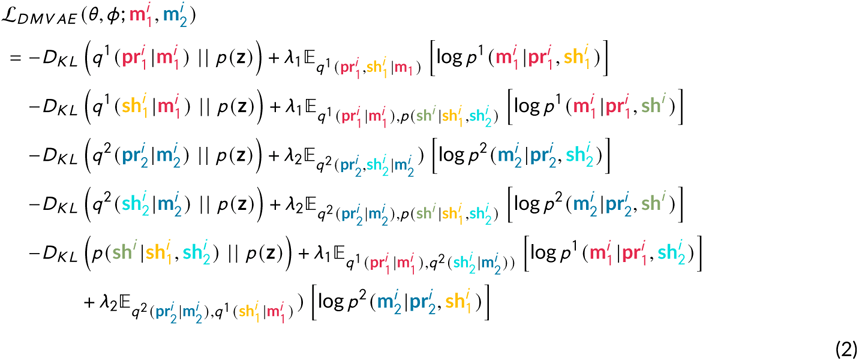

Where *p* (z) is the diagonal Gaussian prior, *λ*_1_ and *λ*_2_ are weighting factors for the reconstruction of each modality, and the last two reconstruction factors are cross-generation factors that use a sample from the prior of their own private space and a sample from the shared space of the other modality for the cross-reconstruction. Once the model has been trained using this objective function, the shared space of the first modality sh_1_ and the second modality sh_2_ should have converged towards a joint shared space sh. Thus, we are left with three separate subspaces; the private space of the first modality pr_1_, the private space of the second modality pr_2_, and the shared space sh. Each subject can be represented as a point in each of these subspaces, this is pictorially represented in Figure 2a. Note that each point, in this case, refers to a normal distribution parameterized by the respective encoder of each modality. The exact implementation of the encoder and decoder for each modality is shown in Figure 1a.

**FIGURE 2.**
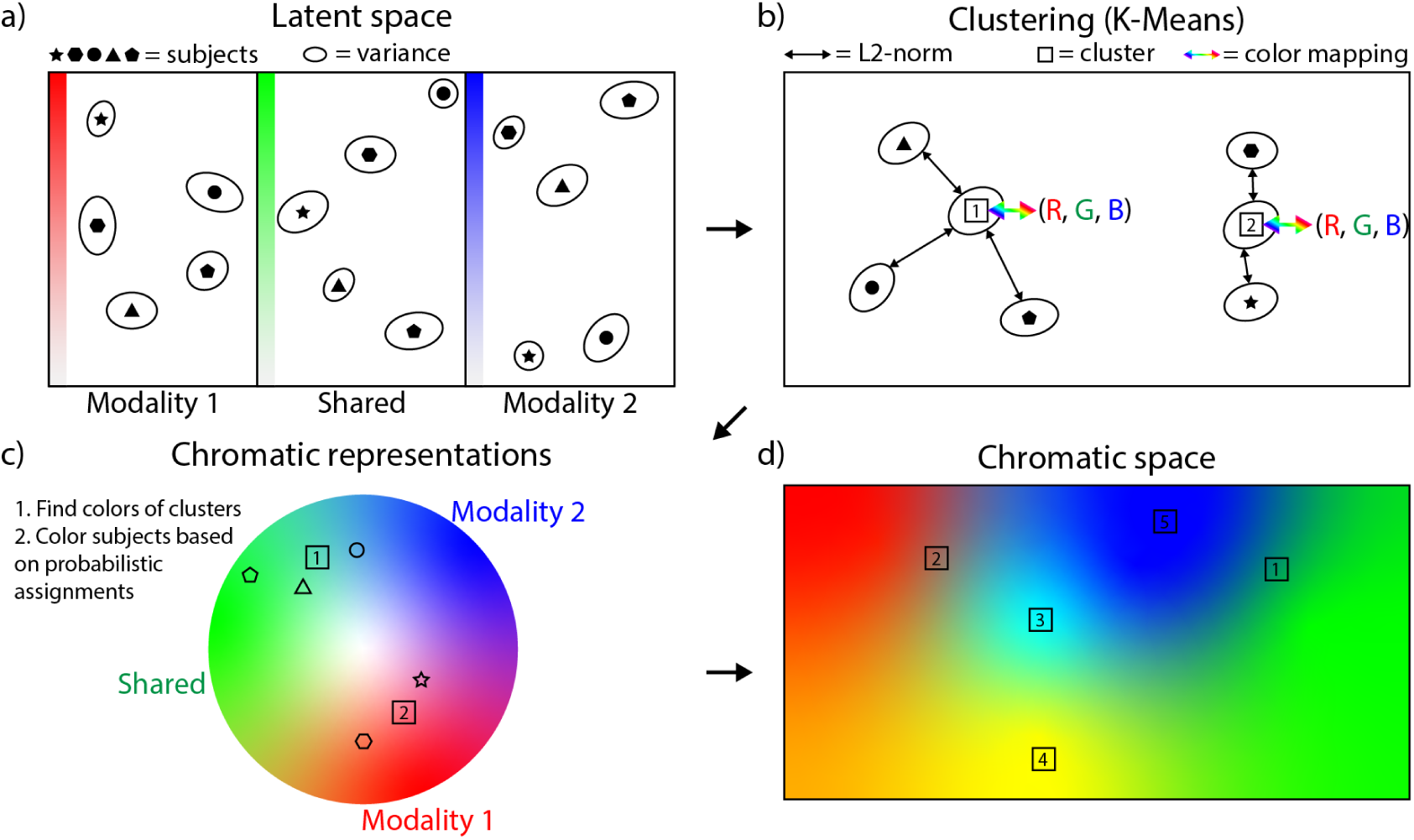
The chromatic fusion framework. This figure shows the steps in the complete chromatic fusion algorithm. The top left shows the latent space that is obtained by training a DMVAE. The top right diagram shows the multidimensional clustering algorithm that is applied to the distributions obtained by the encoder of the DMVAE. Left bottom: the representation of subjects as colors using each meta-chromatic pattern. Right bottom: the final chromatically fused latent space.

### 2.2 Multi-dimensional clustering

After training the model with the objective function in Equation 2, the three subspaces 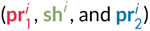 are concatenated together to form one large embedding vector:

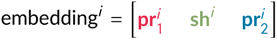

Each subject is represented by its embedding vector, which we use to group the subjects in the dataset for each modality pair. Since each subject is represented by a multivariate normal distribution, we take its most likely value, the mean, to cluster the subjects. The clusters are then found using K-Means++ clustering in the multi-dimensional space. A visualization of the clustering is shown in Figure 2b. To understand the relationship among subspaces and subjects we determine the color of each MCP based on their L2-norm from the mean of the prior distribution, which is zero. The first private space pr_1_ is red, the shared space sh green, and the second private space pr_2_ is blue. How ’red’, ’green’, and ’blue’ an MCP is, is the L2-norm of the cluster in each subspace, divided by the maximum L2-norm for the clusters in that subspace. The norm captures how uncommon a feature is along that specific dimension. We can interpret the norm this way because the neural network is trained by minimizing the KL-divergence between each training subject and the prior during training (see Equation 2). Therefore, high KL-divergence is penalized and the model will try to incur KL-divergence penalties as infrequently as possible. A mean further away from 0 (the mean of the prior), will thus incur a penalty during training. The model will try to ensure a high KL-divergence along a certain latent dimension in the subspace is thus as uncommon as possible. It only uses a high KL-divergence if it aids in the reconstruction error (the other term in the objective function). If not, the magnitude of the KL-divergence would have been optimized to be lower, meaning the subject’s mean deviates less from zero. The ’redness’ of a given MCP can thus be interpreted as patterns that have uncommon features for the first modality. This same interpretation can be applied to the ’greenness’ and ’blueness’ of each MCP. After the decoding process, we can define the full latent space in terms of chromatically fused colors, where subjects are samples from this chromatic space. The colors of each individual subject are determined with a probabilistic assignment to each cluster. This probabilistic assignment is one over the L2 distance to the fourth power between the subject and each cluster, normalized to sum to one. These probabilities are then multiplied by the colors assigned to each MCP. This is pictorially shown in Figure 2c and Figure 2d. We select the number of MCPs for each modality pair based on the elbow criterion, and show this selection process in Appendix B.

### 2.3 Robustness of the multidimensional MCPs

To assess the robustness of the clusters, the dataset is split into 10 stratified folds. Each fold is used as a test set once, and the remaining 9 folds are used to create a training and validation set. The validation set is a random 10% stratified subset of those remaining 9 folds. Validation set subjects are not used to train on, but the objective function (Equation 2) is evaluated on the validation set to ensure the model is not overfitting on the training set. The model with the lowest loss on the validation set is used to cluster the whole dataset. Thus, each cluster occurs 10 times, with different training, validation, and test sets.

We then decode all subjects in our dataset to identify schizophrenia-enriched clusters represented by an MCP in the latent space. In essence, we are trying to establish the robustness of certain subjects being grouped together with respect to changes in the training distribution. To match colors across training folds, we take the clusters in the first fold, fold 0. This fold is also used for further visualizations in subsequent sections (see Section 3). The subjects that are assigned to each cluster in fold 0 are then compared to the subjects assigned to each cluster obtained from training and then clustering on the other 9 folds. The overlap between clusters in fold 0 and the other folds can be expressed as the percentage of subjects that are assigned to both clusters. We use the overlap as the weight in a linear assignment problem and solve the assignment of clusters in fold *k*, *k* ≠ 0 to clusters in fold 0 using the Hungarian algorithm [28, 29]. The average percentage of overlap across all the clusters that were assigned to each cluster in fold 0 indicates the robustness of that cluster with respect to shifts in the training distribution.

The MCPs across the folds are then used to assess specific brain signatures for schizophrenia subjects. By calculating the average percentage of schizophrenia subjects assigned to an MCP across folds, we can understand potential relationships between chromatic colors and subpopulations in the dataset. Given that the model is equipped with a decoder, we can decode schizophrenia-enriched chromatic clusters in the latent space to brain space, and compare them in brain space. The specific hue of the color, as mentioned previously, is a measure of irregularity in terms of the private spaces and the modalities’ shared space. The chromatic colors and interpolations between them are thus a visual and informative measure of specific subpopulations in the dataset.

### 2.4 Understanding heterogeneity in schizophrenia-enriched MCPs

The MCPs we identify capture distinct relationships among each of the modality pairs, but as we hypothesize (Section 1), we expect different modality pairs to partially capture distinct schizophrenia patients. Thus, after clustering the multi-dimensional space for each of the modality pairs, assigning meta-chromatic colors to each of the clusters, and calculating the robustness and percentage of schizophrenia subjects in each MCP, we perform a cross-modality pair analysis. For this analysis, we select schizophrenia-enriched MCPs, at least one for each modality pair, and calculate the percentage of overlap between schizophrenia patients in different clusters, across folds. To perform this analysis across folds, we use the mapping of clusters from all folds to the first fold. Once we obtain the percentage of overlap among schizophrenia-enriched MCPs across modality pairs, we visualize the average overlap across folds normalized by the number of schizophrenia subjects in a particular MCP.

### 2.5 The importance of the shared space

One important aspect of the model we use is the fact it can capture non-linear co-variations between the two modalities in a modality pair. To qualify the assumption that capturing shared information between modalities is essential to understanding complex mental disorders, we visualize shared latent dimensions for two different modality pairs. Additionally, to quantify the importance of the shared space we analyze the stability and correlation to schizophrenia subjects for each of the latent dimensions in a modality pair’s shared space. The results and in-depth analysis are provided in Appendix C. We select an interesting latent dimension from two different modality pairs and show the interpolation from the schizophrenia-enriched part of the latent dimension to the control-enriched part of the latent dimension. Given that the shared space spans multiple dimensions, we can encode the dataset into those dimensions and then calculate the correlation between higher values in a specific dimension and schizophrenia. Since there are no guarantees that a model finds the same latent dimension across each training fold, we only consider latent dimensions that have an average correlation of over 0.7 across folds, see Appendix C.

### 2.6 Cross-reconstruction

Another way to test the shared space is to evaluate how well modalities can be cross-reconstructed from other modalities using the shared space. We can use the shared subspace of the modality that is present plus the prior of the missing modality to create a cross-reconstruction. The modality that is present for a subject, say modality 1, first encodes the data to create a representation that is split into its private subspace pr_1_, and a shared subspace sh_1_. The latter is optimized during training to be similar to the shared subspace of modality two sh_2_. The private subspace of the second modality is then sampled from the prior m_1_. To reconstruct the missing modality, the shared subspace of modality one sh_1_ is concatenated with the private subspace sampled from the prior m_2_ and passed to the decoder. The decoder then reconstructs the sample in the modality’s original space. Note that the prior in this case is the one we trained the DMVAE with, which is a zero-mean unit Gaussian distribution. A complete overview of all the steps we perform in our proposed framework is shown in Figure 3

**FIGURE 3.**
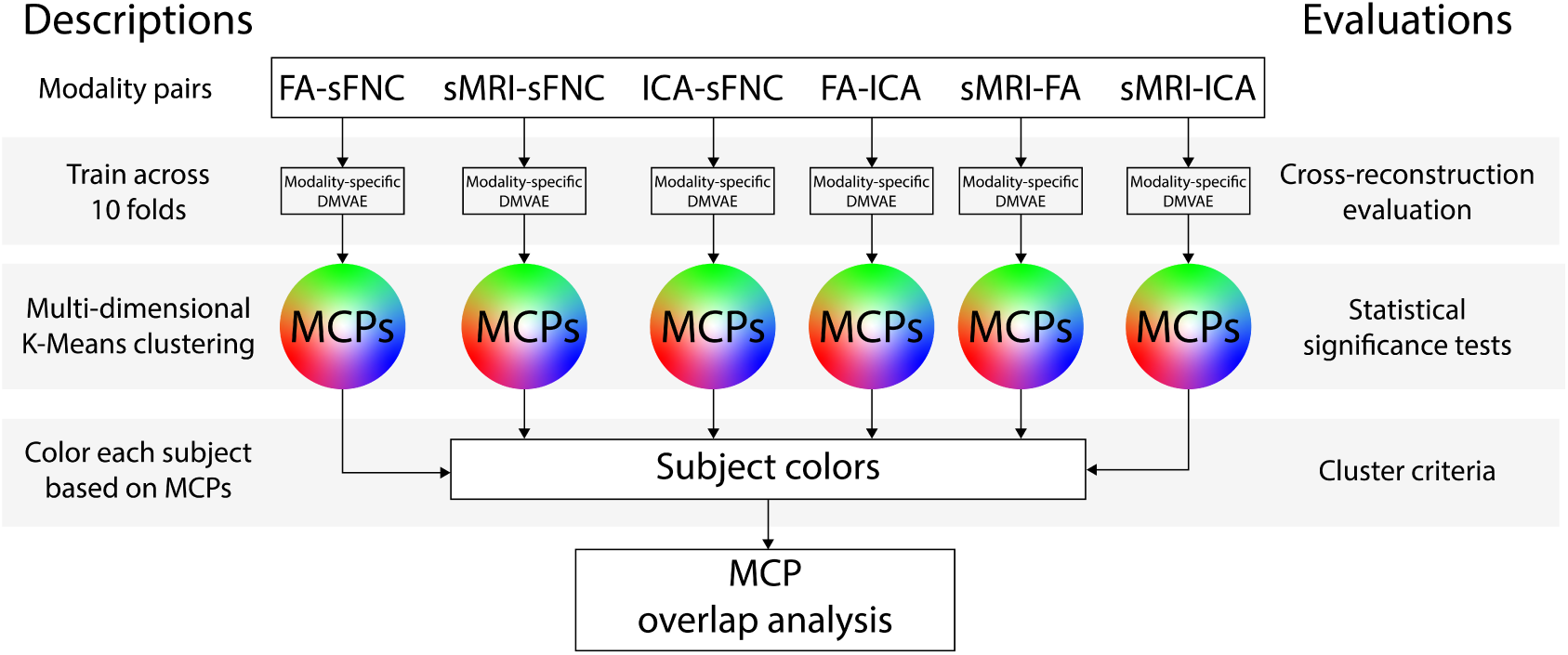
The order of analysis steps used in our chromatic fusion framework. This figure depicts the complete workflow of the analyses we perform in our framework and how they coincide. We start at the top with the modality pairs and work our way down to MCPs, and after that meta-MCPs.

### 2.7 Data

2.7.1 Acquisition and demographics

The main dataset used in this work is the function bioinformatic research network (fBIRN) phase III data, a schizophrenia dataset [30] and the demographics of individuals with each modality pair are described in Table 1. The schizophrenia patients and controls were matched based on age, gender, handedness, and race distributions. Some subjects do not pass the quality assessment of either of the modalities, which means there may be fewer subjects for certain modality pairs. We take the largest number of subjects for each modality pair, such that we maximize the size of the dataset. Interesting in this context is that our method can also generate missing modalities, providing a way to alleviate this issue in the future.

**TABLE 1.** Data sample demographics. Note that P in this case refers to psychosis, AP refers to anti-psychotic medication, and AD refers to anti-depressive medication. Thus, AP and AD in this table refer to the percentage of patients taking anti-psychotic and anti-depressive medication, respectively. PANNs is a symptom scale for schizophrenia, which we have shown split into positive and negative symptoms, and the composite score.

|  | FA-sFNC | sMRI-sFNC | ICA-sFNC | FA-ICA | sMRI-FA | sMRI-ICA |
| --- | --- | --- | --- | --- | --- | --- |
| N subjects | 278 | 311 | 310 | 277 | 278 | 310 |
| Patients (%) | 49.28 | 48.55 | 48.39 | 49.10 | 49.28 | 48.39 |
| Female (%) | 25.54 | 26.05 | 26.13 | 25.63 | 25.54 | 26.13 |
| Female patient (%) | 23.36 | 23.84 | 24.00 | 23.53 | 23.36 | 24.0 |
| Avg Age | 38+-11 | 38+-11 | 38+-11 | 38+-11 | 38+-11 | 38+-11 |
| Avg P length | 17+-12 | 17+-11 | 17+-11 | 17+-12 | 17+-12 | 17+-11 |
| AP (%) | 89.78 | 88.74 | 88.67 | 89.71 | 89.78 | 88.67 |
| AD (%) | 37.23 | 37.09 | 36.67 | 36.76 | 37.23 | 36.67 |
| Avg AP length | 15+-11 | 15+-11 | 15+-11 | 15+-11 | 15+-11 | 15+-11 |
| PANNs positive | 15.5+-5.0 | 15.3+-5.0 | 15.2+-4.8 | 15.4+-4.8 | 15.5+-5.0 | 15.2+-4.8 |
| PANNs negative | 14.5+-5.7 | 14.3+-5.6 | 14.2+-5.6 | 14.4+-5.7 | 14.5+-5.7 | 14.2+-5.6 |
| PANNs composite | -1.1+-6.5 | -1.0+-6.3 | -1.0+-6.3 | -1.0+-6.5 | -1.1+-6.5 | -1.0+-6.3 |

The dataset itself consists of scans collected at seven consortium sites (University of Minnesota, University of Iowa, University of New Mexico, University of North Carolina, University of California Los Angeles, University of California Irvine, and University of California San Francisco). Each consortium records diagnosis, age at the time of the scan, gender, illness duration, symptom scores, and current medication, when available. Furthermore, the inclusion criteria were that participants are between 18 and 65 years of age, and their schizophrenia diagnosis was confirmed by trained raters using the Structured Clinical Interview for DSM-IV (SCID) [31]. All participants with a schizophrenia diagnosis were on a stable dose of antipsychotic medication either typical, atypical, or a combination for at least two months. Each participant with a schizophrenia diagnosis was clinically stable at the time of the scan. The control subjects were excluded based on current or past psychiatric illness based on the SCID assessment or in case a first-degree relative had an Axis-I psychotic disorder. Written informed consent was obtained from all study participants under protocols approved by the Institutional Review Boards at each consortium site.

#### 2.7.2 Preprocessing

We use structural MRI (sMRI), spatial ICA maps, and static functional network connectivity (sFNC) obtained through preprocessing with NeuroMark [32]. The sMRI volumes are preprocessed to voxel-based morphometry (VBM), whereas the spatial ICA maps and sFNC are both obtained by performing ICA on rs-fMRI data using the NeuroMark template. NeuroMark, using the NeuroMark_fMRI_1.0 template (template is released in the GIFT software at http://trendscenter.org/software/gift performs group ICA to extract 53 ICA components from the rs-fMRI signal and calculates the correlation between each to obtain the sFNC. For the spatial ICA components, we selected 8 networks a priori to use in our model to reduce computational complexity, but use all 53 for the sFNC. Each spatial ICA component is used as channels in a 3-dimensional convolutional neural network. Thus, handling more spatial ICA components requires more channels, and more complex training dynamics. The components we use for the spatial ICA components are the supplementary motor area, thalamus, middle inferior frontal gyrus, right inferior frontal gyrus, middle temporal gyrus, precentral gyrus, and inferior frontal gyrus. To obtain the voxel-based morphometry from sMRI data, the data is first processed with SPM 12 in a Matlab 2016 environment. The data is then further processed by segmenting it into modulated gray matter volumes (GMV) and smoothing those segmentations with a 6mm FWHM Gaussian kernel.

For the FA maps, the diffusion-MRI (dMRI) scans were acquired on seven 3T Siemens Tim Trio System and one 3T GE Discovery MR750 scanner at multiple sites. The scanning protocols are described in [30]. The dMRI processing was then performed using FSL (www.fmrib.ox.ac.uk/fsl) and ANTs [33]. The dMRI volumes were first corrected for eddy current distortions and head movement using the eddy (FSL 6.0) with advanced motion-induced signal dropout detection and replacement [34]. Fractional anisotropy (FA) maps were then calculated from the diffusion tensor using dtifit (FSL). FA maps were normalized to the Montreal Neurologic Insititute (MNI) spaced FA template with a nonlinear registration by ANTs. Images with excessive motion, signal dropout, or noise were excluded from further analysis [35, 36].

There are differences in sizes between the FA maps and the sMRI volumes, but we use the same convolutional architecture in the model for each. Namely, the FA maps were sampled at 1mm and the sMRI volumes were sampled at 1.5mm, so we resampled the FA maps to match the sMRI sampling using Scipy [37]. The spatial ICA maps are sampled at 3mm and cropped using a field-of-view (FOV), hence we use a separate convolutional architecture to handle the spatial ICA components, see Figure 1. The mean of the dataset for that modality is removed from each volume, except for the spatial ICA components, where the mean is removed for each subject. We also calculate a group mask for the FA and sMRI maps, to determine which voxels the model should reconstruct. We exclude values below 0.15 after rescaling between [0, 1] for the sMRI and FA data, and exclude values with an absolute value below 0.15 for the spatial ICA maps. Each of the modalities are z-scored based on the mean and standard deviation in the training set.

### 2.8 Experiments

Chromatic fusion is performed on every combination of voxel-based morphometry (VBM), FA maps, static functional connectivity (sFNC), and spatial ICA maps. The model is trained with the AdamP optimizer [38] for 300 epochs with a learning rate of 1*E* − 5. A low learning rate is necessary to ensure stability during training. We use 16 latent dimensions for both private spaces, and 32 dimensions for the shared space for each modality pair. After training a separate model for each modality pair, we identify the MCPs according to the algorithm described in Section 2.2 and evaluate the robustness of each MCP. We visualize the embedding space for each of the modality pairs, and then reconstruct a schizophrenia-enriched MCP in the original space of the modalities for each modality pair. To understand how each of the schizophrenia-enriched MCPs differ across modality pairs, we look at their overlap. A final experiment evaluates the performance of cross-reconstruction for each of the modality pairs on the test folds. The cross-reconstruction is compared to a reconstruction that has access to the private space of the modality (upper bound) and a reconstruction that is created using only the prior distribution (lower bound). To ensure reproducible results, we do not sample from the latent distributions, but rather take their mean because it is the most probable sample under a multivariate normal distribution.

## 3 RESULTS

### 3.1 MCP analysis

To quantify whether an MCP potentially represents neuropsychiatric and/or demographic factors we calculate the percentage of subjects in an MCP that also belong to a stratum. These percentages for sex and schizophrenia diagnosis are shown in Table 2.

**TABLE 2.** The meta-chromatic patterns for each modality pair. We show the percentage-wise robustness (-R), subjects who are female (-F), and subjects diagnosed with schizophrenia (-SZ) for each MCP. The text of some schizophrenia-enriched MCPs (percentage above 70%) is made bold. Each cell’s color indicates the color of the corresponding MCP, based on chromatic fusion.

| MCPs | FA-sFNC | sMRI-sFNC | ICA-sFNC | FA-ICA | sMRI-FA | sMRI-ICA |
| --- | --- | --- | --- | --- | --- | --- |
| 0-R | 74+-11 | 78+-14 | 96+-2 | 44+-12 | 65+-18 | 52+-18 |
| 0-SZ | 75+-15 | 48+-12 | 62+-1 | 47+-12 | 27+-7 | 50+-11 |
| 0-F | 29+-5 | 30+-5 | 23+-1 | 31+-4 | 40+-7 | 24+-6 |
| 1-R | 75+-12 | 78+-11 | 97+-2 | 72+-13 | 75+-12 | 49+-14 |
| 1-SZ | 28+-3 | **70+-13** | 23+-1 | **73+-4** | **72+-5** | 47+-10 |
| 1-F | 20+-4 | 24+-4 | 27+-1 | 24+-3 | 18+-2 | 21+-7 |
| 2-R | 54+-22 | 82+-12 | 87+-6 | 66+-14 | 70+-12 | 43+-14 |
| 2-SZ | 28+-3 | 29+-3 | **77+-2** | 35+-6 | 55+-10 | 67+-6 |
| 2-F | 20+-4 | 25+-2 | 31+-2 | 19+-5 | 27+-7 | 18+-7 |
| 3-R | 62+-16 | - | 87+-5 | 52+-17 | 72+-17 | 71+-13 |
| 3-SZ | **70+-10** | - | 29+-2 | 53+-5 | 50+-4 | 20+-4 |
| 3-F | 29+-3 | - | 25+-2 | 29+-3 | 17+-5 | 38+-3 |
| 4-R | - | - | - | 93+-13 | - | 63+-17 |
| 4-SZ | - | - | - | **84+-3** | - | **78+-5** |
| 4-F | - | - | - | 20+-4 | - | 15+-6 |
| 5-R | - | - | - | 31+-18 | - | 46+-18 |
| 5-SZ | - | - | - | 50+-19 | - | 44+-13 |
| 5-F | - | - | - | 25+-6 | - | 34+-7 |

The results in Table 2 show there is an interesting combination between unstable MCPs and more stable MCPs with fairly high percentages of subgroups in the population. All modality pairs produce at least one or two interesting MCPs, even if those MCPs are not as robust. To test whether the MCP pairs were significant in terms of the percentage of schizophrenia or female subjects captured by the MCP, we performed significance analyses, see Appendix A. Since our model captures non-linear relationships, we cannot simply regress out site effects beforehand, which is why we chose to assess post-hoc if there were significant site effects for any of the MCPs with respect to the percentage of schizophrenia patients in a cluster. To do this, we calculated the standard deviation between the values shown in Table 2, and the median percentage of schizophrenia patients in a cluster over folds, and for each site. The standard deviation is less than 5% for all MCPs, and often around 1-2%. This indicates that the percentage of patients in each MCP is generally stable across sites.

Every single modality pair also includes at least one MCP enriched for schizophrenia vs controls (more than 70% schizophrenia patients in the MCP), these MCPs are highlighted as bold in Table 2. Importantly, most of these schizophrenia-enriched clusters have low standard deviations across folds, and are often also robust. For example, for the ICA-sFNC pair, MCP 2 is highly robust (87%) with 77% schizophrenia subjects, and is green, meaning the irregularities are especially large in the shared space. For the FA-sFNC and sMRI-FA pairs, the most schizophrenia-enriched MCPs (3, and 1, respectively) are red-ish and blue, respectively. Since for the FA-sFNC pair, the FA maps are the first private space, and for the sMRI-FA pair, the FA maps constitute the second private space, these MCPs both indicate irregularities in the FA maps for the schizophrenia-enriched MCPs. However, for FA-ICA and sMRI-ICA, cluster 4 contains irregularities in all subspaces, fusing into a white color. Additionally, sMRI-sFNC MCP 1 is purple, which means that the irregularities mostly occur in both private subspaces (red and blue). Importantly, it is notable that none of the schizophrenia-enriched MCPs are particularly skewed in terms of the number of male/female subjects, compared to the percentage of female subjects in the dataset (see Table 1).

The importance of the schizophrenia-enriched MCPs is also apparent from the significance analysis in Appendix A. None of the clusters have significant differences in subjects based on sex, except for the sMRI-FA modality pair. MCP 3 for the FA-sFNC pair contains a corrected significant number of schizophrenia subjects compared to all other MCPs 0 (p=0.017), 1 (p< 0.0005), and 2 (p=0.039). Similarly, for MCP 1 in the sMRI-sFNC pair: MCP 0 (p< 0.0005) and MCP 2 (p< 0.0005). MCP 2 in the ICA-sFNC pair contains a corrected significant number of schizophrenia compared to MCPs 1 (p< 0.0005) and 3 (p< 0.0005), but not 0. This is because MCP 0 also contains a relatively large number of schizophrenia subjects (62%), which is significant compared to MCPs 1 (p< 0.0005) and 3 (p< 0.0005). Both FA-ICA MCP 1 and 4 are enriched for schizophrenia subjects, but only MCP 1 contains a significant number of subjects compared to all other clusters (except 4), namely 0 (p=0.001), 2 (p< 0.0005), 3 (p=0.037), and 5 (p=0.003). This is due to the low number of subjects in cluster 4, and hence after correction, the cluster is not significantly different in terms of the number of schizophrenia subjects from other clusters. However, the cluster itself is robust across folds 93%, and contains 84% schizophrenia subjects, so we do include this cluster in further analyses. For the sMRI-FA pair, MCP 1 contains a significant number of schizophrenia subjects compared to clusters 0 (p< 0.0005) and 3 (p= 0.042). MCP 0 for this modality pair actually contains a significantly small number of schizophrenia subjects compared to all other MCPs: 1 (p< 0.0005), 2 (p< 0.0005), and 3 (p= 0.002). For the sMRI-ICA pair, MCP 4 contains a significant number of schizophrenia subjects compared only to MCP 3 (p=< 0.0005). This is because all other clusters contain roughly 50% schizophrenia subjects. For this modality pair, the schizophrenia subjects thus appear especially heterogeneous.

### 3.2 Chromatic fusion space shapes

We visualize the embedding space for each modality pair in Figure 4. The colors of the MCPs and subjects in the figure are based on the L2-norm from a subject to its assigned cluster, as explained in Section 2.2. To visualize the 64-dimensional subjects and clusters, the mean of the distributions are visualized in a 2-dimensional space using t-SNE [39].

**FIGURE 4.**
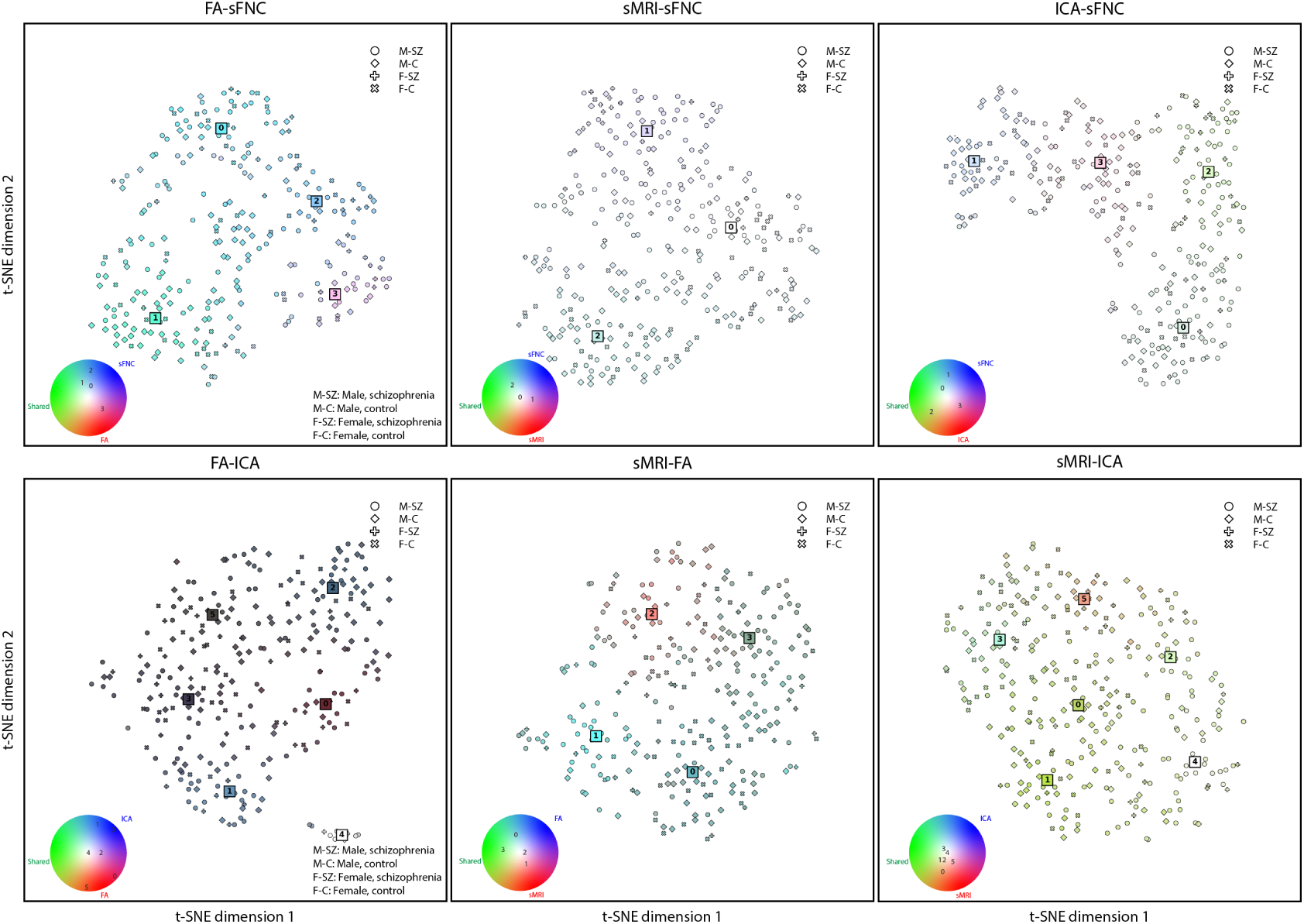
At-SNE plot of the embedding space for each of the modality pairs. The color wheels in the bottom left of each subfigure show the location of MCPs on the color wheel in terms of their irregularities in the shared and private subspaces.

Each modality pair clearly has unique color patterns and structures in the embedding space, shown in Figure 4. Furthermore, this visualization highlights how MCPs roughly compare to each other spatially in the embedded chromatic space. It is clear from this view that MCP 4 in the FA-ICA is a small MCP with very irregular subjects (white color), seemingly far away from the rest of the MCPs. This leads to very dark colors for the other MCPs because their irregularities are not as extreme as this MCP. In this case, we have put the MCPs in the color wheel legend near the color that they are closest to perceptually and numerically since the color wheel does not contain dark colors. Additionally, for MCP 1 and MCP 2 the FA-sFNC and sMRI-sFNC modality pairs, respectively, seem to be further away from the other MCPs. Neither are schizophrenia-enriched MCPs, but rather control-enriched MCPs. For the sMRI-ICA modality pair, however, it is harder to make clear distinctions, which is also reflected in the colors of the MCPs and how chromatic the space is. For this modality pair, most MCPs have similar colors, and as a result, their subjects are also colored a reddish green.

### 3.3 Visualizing schizophrenia-enriched MCPs

To qualitatively understand what types of irregularities the main schizophrenia-enriched MCPs represent, we have reconstructed each MCP using the decoder in our model. Instead of using the MCP center however, we take the schizophrenia patients assigned to each schizophrenia-enriched MCP and reconstruct their average in the latent space. This allows us to directly look at the average of the schizophrenia subjects, without adding heterogeneity from control subjects. Figure 5 shows these reconstructions for each modality pair.

**FIGURE 5.**
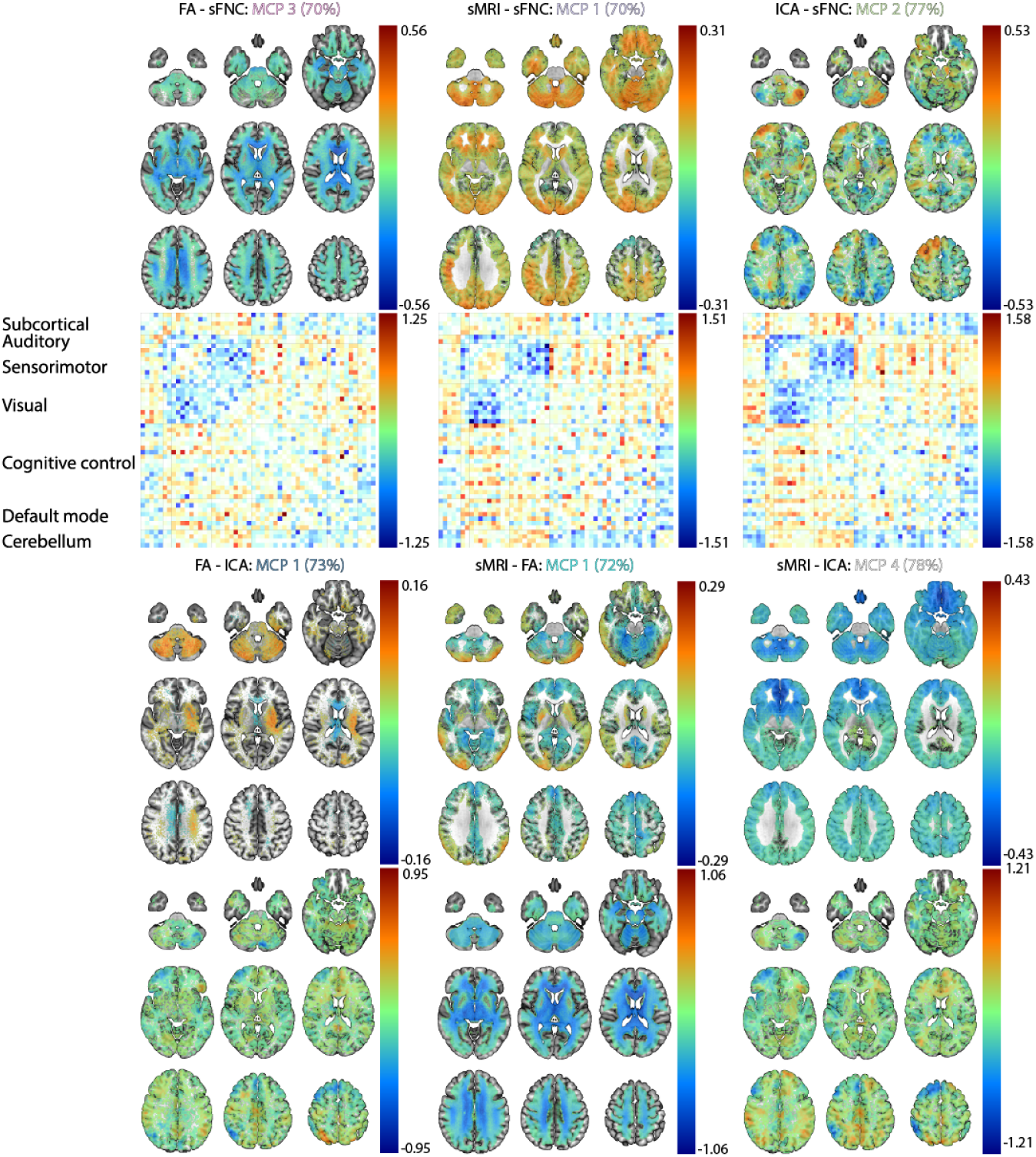
The reconstruction of schizophrenia-enriched MCPs. The percentages next to the name of the MCP indicate the average percentage of schizophrenia subjects in the cluster across folds. Note that the values for the color bar differ for each modality pair to ensure there is enough contrast to see differences in positive and negative changes.

The reconstructions of the MCPs in Figure 5 show interesting patterns. Mainly, we observe differences between MCPs that have the same modalities. For instance, the sMRI patterns in the schizophrenia-enriched MCP for the sMRI-sFNC modality pair has opposite directionality in some areas compared to the schizophrenia-enriched sMRI-FA and sMRI-ICA pairs. A similar pattern of opposite directionality occurs for the spatial ICA patterns for the ICA-sFNC MCP, when compared with FA-ICA and sMRI-ICA. To test our hypothesis that these MCPs capture different subgroups of schizophrenia pattients, we perform a heterogeneity analysis on the schizophrenia-enriched MCPs.

### 3.4 Heterogeneity of schizophrenia as a function of modality pair

The fact that different modality pairs lead to schizophrenia-enriched clusters with different and sometimes even opposing modality-based differences ties into our next results. Namely, how many schizophrenia subjects are distinctly captured by each of the schizophrenia-enriched MCPs. For this analysis, we use all schizophrenia-enriched MCPs (%SZ > 70): FA-sFNC-MCP-3, sMRI-sFNC-MCP-1, ICA-sFNC-MCP-2, FA-ICA-MCP-1, and FA-ICA-MCP-4, sMRI-FA-MCP-1, and sMRI-ICA-MCP-1. On average across folds, these clusters account for 65%+-4 of all schizophrenia subjects in the dataset. The results in Figure 6 show the average percentage of schizophrenia subjects captured by the row-wise MCP not captured by the column-wise MCP, divided by the number of schizophrenia subjects in the row-wise MCP. This is thus a metric of distinct schizophrenia subjects captured by the row-wise MCP, and a higher percentage means the MCP captures more distinct schizophrenia subjects. The MCPs are sorted based on the percentage each schizophrenia-enriched MCP captures on its own, out of all schizophrenia subjects they capture together. This percentage is shown on the diagonal for each MCP.

**FIGURE 6.**
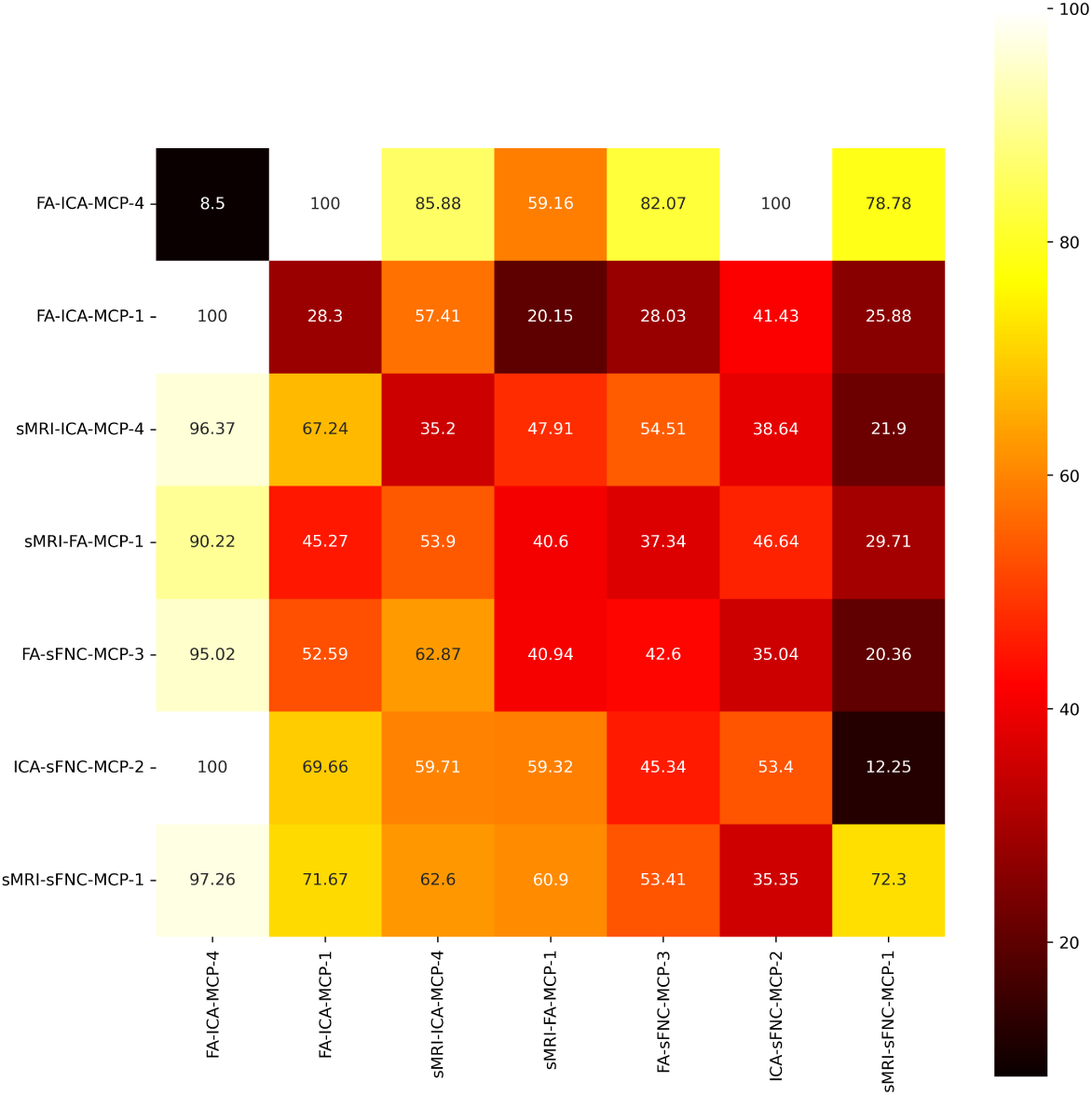
The distinct percentage of schizophrenia subjects each of the schizophrenia-enriched MCPs capture. The percentages are normalized by the number of subjects in the row MCP. The MCPs are sorted based on the percentage each schizophrenia-enriched MCP captures on its own, out of all schizophrenia subjects they capture together. This percentage is shown on the diagonal for each MCP.

The most distinct aspect of Figure 6 is that all MCPs in the first column capture almost fully distinct schizophrenia subjects. This is likely because FA-ICA-MCP-4 only captures 8.5% of all schizophrenia subjects captured by all of these MCPs together. In fact, looking at the first row, we can see that the schizophrenia subjects captured by FA-ICA-MCP-4 are almost never captured by other MCPs, except for sMRI-FA-MCP-1, but never fully. Especially distinct MCPs are ones with high values above the diagonal because that means the MCP contains many distinct schizophrenia subjects when compared with a larger MCP than itself. Specifically, FA-ICA-MCP-1, sMRI-ICA-4, and sMRI-FA-MCP-1 contain many distinct schizophrenia subjects when compared with sMRI-ICA-4, FA-sFNC-MCP-3, and sMRI-sFNC-MCP-1, respectively. Notably, these clusters all do not contain sFNC and all the largest clusters do contain sFNC. This potentially indicates that heterogeneity, and clear subgroups, are a function of what modality pairs are combined to analyze schizophrenia.

### 3.5 The importance of the shared space

We perform a more in-depth analysis of the robustness across folds and correlation of each latent dimension in the shared space to schizophrenia in Appendix C. Based on the most robust shared latent dimensions, we select two latent dimensions to highlight how interpolating from one end of the dimension to the other leads to covariations of the modality pair. For the following latent dimensions these interpolations are aligned with how schizophrenia diagnosis and they are shown in Figure 7.

**FIGURE 7.**
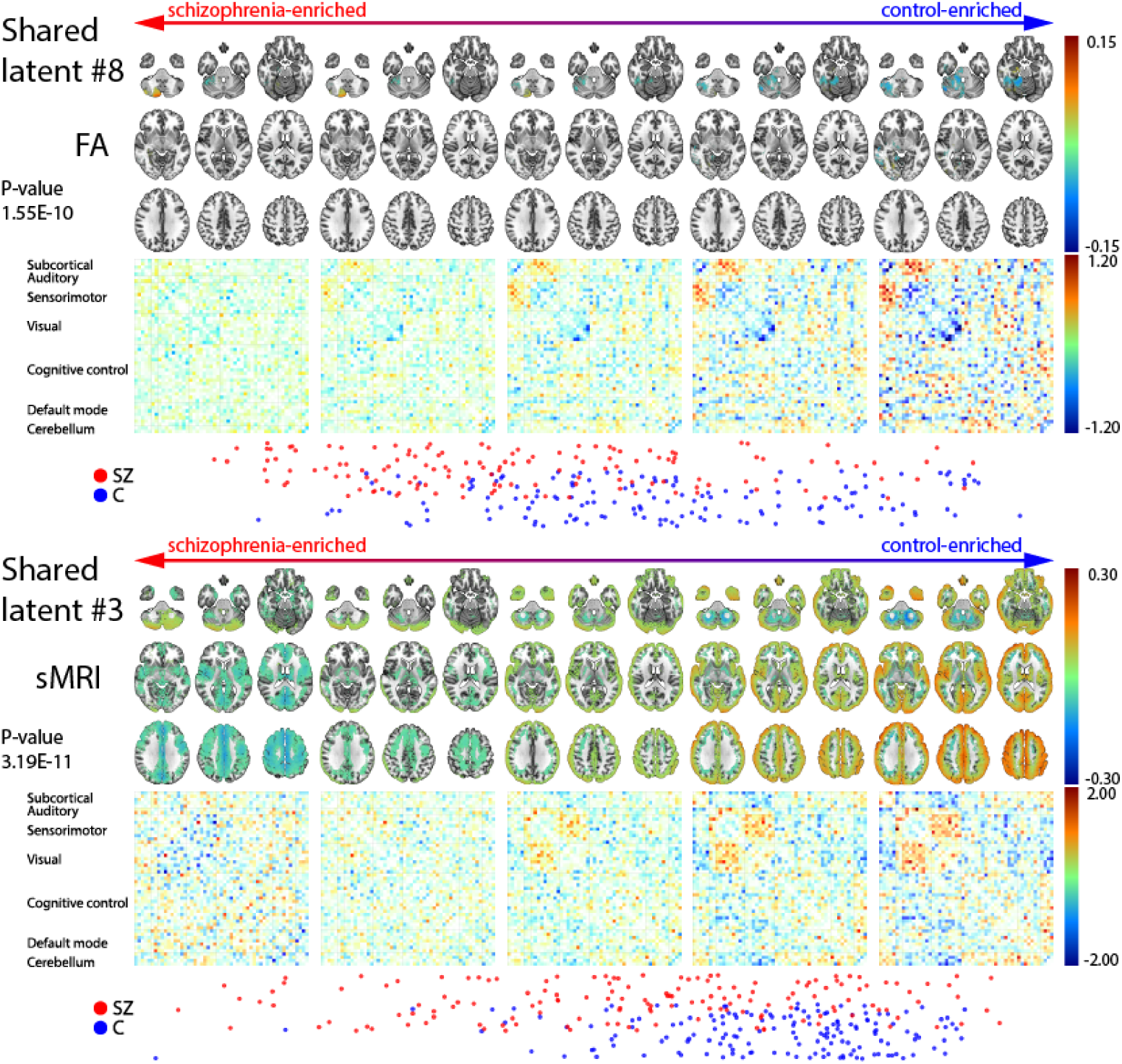
Interpolations for two representative latent dimensions from schizophrenia-enriched to control-enriched. The two latent dimensions are taken from the FA-sFNC, and sMRI-sFNC modality pairs, and show how the modalities change along the latent dimension, from a more schizophrenia-enriched part of the subspace to a control-enriched part of the subspace. We have included a plot of the schizophrenia and control subjects, and the p-value for fold 0 in the figure.

Figure 7 indicates that in the FA-sFNC pair, as we interpolate from schizophrenia subjects to control subjects, we see an increase in modularity and connectivity between the subcortical and sensorimotor regions, and a decrease in self-connectivity for the visual regions. This is coupled with higher fractional anisotropy in the cerebellum for schizophrenia subjects that non-linearly interpolates into a decrease in fractional anisotropy in a more anterior part of the cerebellum. Since this is a shared space, it is interesting that the interpolations for both modalities follow a gradient for the cerebellum. For the sMRI-sFNC pair, we almost see a split in the scatter plot at the bottom of Figure 7 (around the fourth panel from the left) between schizophrenia patients with higher and lower general voxel-based morphometry. Although the voxel-based morphometry in the cerebellum for more schizophrenia-enriched patients interpolates to a decrease in voxel-based morphometry and moves more anterior, the rest of the brain moves from lower voxel-based morphometry to increased voxel-based morphometry on the control-enriched side. This is coupled with an increase in the modularity of the sFNC, but also an increase in the connection between sensorimotor and visual regions for control subjects.

### 3.6 Cross-reconstruction

To evaluate the shared space with respect to how well it can help reconstruct other modalities, we compare the performance of the cross-reconstructions across all the modality pairs. The metric we use to compare the cross-reconstruction with two baselines is the mean squared error of the reconstruction. The difference is evaluated on the test set and averaged across all 10 folds, and the results are shown in Table 3. Cross-reconstruction is evaluated for two baselines. The first baseline uses the mean of the prior to reconstruct the ’missing’ modality. The second baseline is called normal and uses the mean of the distribution predicted by the encoder to reconstruct the modality. The cross-reconstruction uses the shared subspace of the other modality and the mean of the prior of the private space to reconstruct the ’missing’ modality. The mean of the prior for both the prior baseline and cross-reconstruction results is zero.

**TABLE 3.** The mean squared error for cross-reconstructed modalities. The mean squared error is calculated between the voxels of the reconstructed volumes and the ground truth volumes.

| Modalities | Prior | Normal | Cross |
| --- | --- | --- | --- |
| sMRI → FA | 0.9902+-0.2155 | 0.6814+-0.1278 | 0.7108+-0.1411 |
| FA → sMRI | 0.9717+-0.0451 | 0.8681+-0.0344 | 0.9219+-0.0444 |
| FA → sFNC | 0.9717+-0.0451 | 0.8761+-0.0357 | 0.9113+-0.0369 |
| sFNC → FA | 1.0245+-0.0460 | 0.8197+-0.0324 | 1.0152+-0.0431 |
| sMRI → sFNC | 0.9874+-0.1635 | 0.7012+-0.0942 | 0.7683+-0.1159 |
| sFNC → sMRI | 1.0198+-0.0659 | 0.8091+-0.0464 | 1.0022+-0.0624 |
| ICA → sFNC | 1.0000+-0.0056 | 0.9960+-0.0056 | 0.9975+-0.0057 |
| sFNC → ICA | 1.0239+-0.0532 | 0.7995+-0.0308 | 0.9212+-0.0417 |
| FA → ICA | 0.9709+-0.0828 | 0.8646+-0.0433 | 0.8706+-0.0459 |
| ICA → FA | 1.0001+-0.0067 | 0.9983+-0.0073 | 1.0012+-0.0060 |
| sMRI → ICA | 0.9878+-0.1252 | 0.6747+-0.0727 | 0.6938+-0.0798 |
| ICA → sMRI | 1.0000+-0.0056 | 0.9972+-0.0057 | 1.0000+-0.0055 |

Table 3 shows that the cross-reconstruction of modalities generally improves with the shared information. Although the results differ in terms of the magnitude of the improvement, all of these pairs have a fairly high KL-divergence in the shared space, which indicates information encoded in that shared space. Thus, these results indicate that the shared information generally helps reconstruction, sometimes even close to knowing the private space of the other modality, but that how much it helps depends on what modality cross-reconstructs which.

## 4 DISCUSSION

In this work, we present an intuitive and flexible framework to facilitate new insights into multimodal neuroimaging data. By representing information from modalities as colors, we can intuitively visualize the contribution of modalities along dimensional relationships. Our method identifies schizophrenia-enriched clusters for each modality pair. The clusters are assigned colors to indicate important meta-chromatic patterns (MCPs) and define a chromatic space. Although these MCPs overlap across modality pairs in terms of what subjects are captured by schizophrenia-enriched MCPs, we find that different modality pairs highlight distinct subgroups of schizophrenia subjects. Together with visualizations and statistical results for the schizophrenia-enriched MCPs, and interpolations of the shared dimensions for certain modality pairs, we show how critical multi-modal analyses are to further our understanding of spectrum psychiatric disorders, such as schizophrenia.

Our analyses are centered around a framework that views the contributions of modalities in modality pairs as colors. Especially as we move to the inclusion of more modalities, the visualization, and interpretation of combinations of modalities are more naturally done by considering them as different colored lights that chromatically fuse into a certain perspective on diseases or demographic variables. Furthermore, to keep in line with the RDoC initiative, we propose a framework that allows multiple modalities or units of analysis to be included in a single framework that aims to study a mental disorder. We refrain from binary labels and only use unsupervised methods to study schizophrenia, and find that the schizophrenia-enriched MCPs capture more than 65% of the schizophrenia subjects in our sample. This means that along either of the modality pairs, these schizophrenia subjects show clear deviations from the rest of the sample, which is subsequently captured in these schizophrenia-enriched MCPs. To dive deeper into what these specific deviations mean in the original space of the modality, we visualize the most important schizophrenia-enriched MCP for each modality pair, revealing results that align with previous work, but also extend it in an interesting, and understudied multimodal way.

Our findings regarding schizophrenia-enriched meta-chromatic patterns in Figure 5 show some general trends. Firstly, each of the schizophrenia-enriched MCPs for the modality pairs that contain sFNC in its pair, shows hypoconnectivity between visual and sensorimotor regions and especially for the ICA-sFNC pair hyperconnectivity between visual-cerebellum and visual-subcortical regions. This is a general trend in each of the MCPs, although which components are most hypo-connected and how hyperconnected the aforementioned regions are differs for each of the modality pairs. Hypoconnectivity between the visual and sensorimotor cortex for schizophrenia subjects has pre-viously been linked to schizophrenia [40]. Reduced connectivity between these areas may have an impact on self-processing in patients [40] and generally be related to early-stage visual processing deficits in schizophrenia [41]. Furthermore, subcortical-visual and cerebellum-visual hyperconnectivity aligns with previous unimodal work within this cohort [42, 43] and in other cohorts [44]. These connectivity patterns are paired with reduced fractional anisotropy near the corpus callosum and regions superior to the corpus callosum in the FA-sFNC pair. Notably, a decrease in FA strength in the cerebellum, corpus callosum, and superior longitudinal fasciculi have previously been linked to schizophrenia [45, 46]. The decrease in fractional anisotropy especially near the corpus callosum is potentially related to schizophrenia-like psychosis due to structural defects [45, 47]. We see similar patterns of reduced FA strength in the sMRI-FA pair, where decreased FA is also more prominent in the cerebellum. Interestingly, the FA-ICA pair shows increased FA strength in the cerebellum, although only slightly. Potentially reflecting a slightly distinct schizophrenia subgroup without the reduced FA strength in the cerebellum. For the sMRI-sFNC pair, the connectivity pattern for the sFNC are linked with increased voxel-based morphometry most pronounced in the cerebellum, occipital lobe, and near the motor areas, with some reduced voxel-based morphometry in the superior frontal lobe. For all other sMRI-based MCPs, we generally see reductions in frontal lobe voxel-based morphometry, which is in line with previous work [48]. In a similar opposite directional pattern, the sMRI-sFNC MCP may capture some schizophrenia subjects with different patterns from the other sMRI-based MCPs, accentuating schizophrenia’s heterogeneity. We also observe a different pattern for the ICA-based MCPs, where the ICA-sFNC shows increased spatial ICA map strength in the left frontal lobe and directional asymmetry in the cerebellum (decrease on the left and increase on the right). However, for the FA-ICA and sMRI-ICA MCPs, the spatial ICA maps have decreased spatial ICA map strengths in the left frontal lobe. For ICA-sFNC and sMRI-sFNC in Figure 6, we also observe that these MCPs capture a fairly large percentage of unique schizophrenia subjects compared to other MCPs that are also ICA and sMRI-based, respectively. Together, the results in Figure 5 and Figure 6 clearly indicate that our method can find interesting schizophrenia subgroups, indicating that heterogeneity may be a function of what modalities are paired during training.

An important aspect of schizophrenia we highlight in this work is how the meta-chromatic patterns we find are associated with a different subgroup of schizophrenia patients in the sample. By analyzing a wider range of modalities, and specifically training our model to find interpretable spaces between pairs of modalities, we are able to understand how these subgroups change with different modality pairs. Although the subgroups are not unique for each modality pair, it is clear that additional modalities provide us with information about subjects that may not have been captured by a schizophrenia-enriched cluster if we had only considered one or two modalities. This also speaks to the heterogeneous nature of schizophrenia, where some subjects may deviate across most brain measures, and some subjects only deviate along specific brain measures or the shared information in specific brain measures. By highlighting schizophrenia from multiple directions, and conceptualizing an intuitive framework around this type of analysis, we also specifically follow the trans-diagnostic NIMH research domain criteria (RDoC) initiative [7].

We also emphasize another important finding regarding spectrum mental disorders: the significance of extracting shared information from a pair of modalities. In Appendix C, we demonstrate that for each modality pair, there exist replicable shared latent dimensions. Furthermore, we observe that at least a few of these latent dimensions exhibit high correlations to schizophrenia. The reason that these latent dimensions are of particular interest is that we can easily interpolate between ends of the dimension that are either control-enriched or schizophrenia-enriched. We also find that the interpolations are largely non-linear and could not have been captured by a linear model, which are still common. These interpolations help us view certain normative imaging deviations related to schizophrenia on a spectrum as well. There is a wealth of information that is encoded exactly on the border between schizophrenia-enriched and control-enriched subspaces, namely how the extracted shared features transition from schizophrenia to controls. Specifically, the significance of the correlations to schizophrenia as shown in Figure 7 highlight its potential for future analyses. The patterns we see change along two highlighted latent dimensions show distinct modality-specific patterns exist at the extreme end of the interpolative patterns. Furthermore, we see evidence of reduced modular functional patterns on the schizophrenia-enriched side of the latent dimension, as linked to schizophrenia previously [49], and focal spatial regions, for the FA-sFNC pair specifically. Notably, the patterns we obtain for sFNC, which is part of both modality pairs, is different for both the highlighted FA-sFNC and sMRI-sFNC latent dimension. This again indicates how coupling different modalities can result in distinct patterns being uncovered by multimodal machine learning models.

### 4.1 Limitations

The validity of meta-chromatic patterns is partly dependent on the quality of the reconstructions that the DMVAE model produces. It is therefore important to improve the reconstructions by adapting the model or architectures to neuroimaging modalities and improve the cross-reconstructions between the modalities. Furthermore, not all of the MCPs are as robust as others, which is likely due to the stochastic nature of training a variational autoencoder and the differences in distribution between each of the folds. This can be improved by imposing more inductive biases into the architecture and model, or by using larger datasets. This leads to another point, namely that this framework needs to be tested on more datasets to show its robustness across datasets.

It is challenging to address site effects in the context of machine learning models that can capture complex and nonlinear relationships. To evaluate the potential impact of the acquisition site on the result we calculated the standard deviation between the median (across folds) percentage of patients from each site in a cluster. Results showed that the percentage of patients in a cluster was always within 5% of the mean. The patient/control ratio was thus consistent across sites. This gives us confidence in the robustness of the results to site effects. However, for larger datasets analyzed in future work, we plan to continue to evaluate this issue and develop additional metrics that can quantify site effects.

Other than site effects, schizophrenia clusters could also be proxies for uncontrolled confounders, such as severe mental illness, prolonged exposure to psychotropic drugs, and lower socioeconomic status. This is a more general issue with data-driven schizophrenia analysis and should be carefully assessed in exploratory studies.

### 4.2 Future work

The framework can be expanded to additional datasets and more modalities. Future work can also extend the model and MCP framework to move beyond pair-wise to N-way unique and shared links among modalities. Additionally, there is a potential to discover more entangled shared factors by learning representations from minimally pre-processed rs-fMRI directly, together with the FA maps and structural MRI volumes. The incorporation of rs-fMRI as a timeseries, rather than static FNC will facilitate additional insights as in parallel group ICA + ICA [50] and also allow for the fusion of other dynamic modalities, such as EEG. Learning representations from unprocessed modalities can be coupled with ingenious ways to incorporate inductive biases such as group differentiation, regularizations on the weights in the network, or constraints on the latent factors such as sparsity. There are thus wide-ranging possibilities to increase the utility and generality of this framework.

## 5 CONCLUSION

The presented framework can be used in various ways, both as an exploratory tool, to perform hypothesis-testing, to evaluate the enrichment of modalities with each other, and potential specific inter-modality patterns predicted apriori, or to evaluate the heterogeneity of schizophrenia subjects within a sample. Our model can visualize individualized inter-modal relationships, a novel aspect of our work relevant to many applications, including brain development/aging, clinical studies, etc. To conclude, we have shown this framework mostly as an exploratory tool but linked our findings back to previous work on schizophrenia or on this sample specifically. In doing this, we showed how training an autoencoder to find private and shared spaces from a pair of modalities leads to meta-chromatic patterns that are schizophrenia-enriched, each modality pair with a different schizophrenia subgroup. We also take a first step towards understanding and evaluating the heterogeneity among schizophrenia subjects in our sample and visualize interesting inter-modality patterns and interpolations. For instance, we observe a decrease in the modularity of functional connectivity and decreased visual-sensorimotor connectivity for schizophrenia patients for the FA-sFNC and sMRI-sFNC modality pairs, respectively. The visual-sensorimotor hypoconnectivity may indicate impaired self-processing in patients. Additionally, our results generally indicate decreased fractional corpus callosum anisotropy, which is linked to psychosis, and decreased spatial ICA map and voxel-based morphometry strength in the superior frontal lobe for patients across multiple modality pairs.

## Data Availability

All data in the present study can be obtained from the original authors of the dataset. All results in the present study are available upon request to the authors.

## Acknowledgements

This material is supported by the National Science Foundation under Grant No. 2112455 and the National Institutes of Health grant #R01EB006841. Eloy Geenjaar was supported by the Georgia Tech/Emory NIH/NIBIB Training Program in Computational Neural-engineering (T32EB025816).

## Data Availability

Data sharing is not applicable to this article as no new data were created or analyzed in this study.

## A MCP STRATA SIGNIFICANCE

The following subsections report the p-values for cluster pairs and how significantly one cluster codes for schizophrenia or sex more than another. To calculate the p-values, we loop over all of the cluster pairs and obtain all of the subjects that are in the two clusters for each fold. We first assign subjects to the cluster that it is most commonly assigned to across folds, and in case a cluster is assigned the most to two different clusters (e.g. cluster 1 in 5 folds and cluster 2 in the other 5 folds), we assign the subject to the cluster it was assigned to in fold 0. We do this because we use fold 0 for visualizations. Then we calculate the Pearson correlation between the assigned cluster labels (0 or 1) and a subject’s schizophrenia diagnosis, and between the cluster label and a subject’s sex. After calculating the p-value for each modality pair we perform Bonferroni correction across each cluster pair for a cluster. The results are shown in subsections for each modality pair below.

### A.1 FA-sFNC

**TABLE 4.** The significance for the sex stratum

| Cluster pairs | C0 | C1 | C2 | C3 |
| --- | --- | --- | --- | --- |
| C0 | NA | 0.562 | 1.0 | 1.0 |
| C1 | 0.562 | NA | 1.0 | 1.0 |
| C2 | 1.0 | 1.0 | NA | 1.0 |
| C3 | 1.0 | 1.0 | 1.0 | NA |

**TABLE 5.** The significance for the schizophrenia stratum

| Cluster pairs | C0 | C1 | C2 | C3 |
| --- | --- | --- | --- | --- |
| C0 | NA | 0.002 | 1.0 | 0.017 |
| C1 | 0.002 | NA | 0.009 | <0.0005 |
| C2 | 1.0 | 0.009 | NA | 0.039 |
| C3 | 0.017 | <0.0005 | 0.039 | NA |

### A.2 sMRI-sFNC

**TABLE 6.** The significance for the sex stratum

| Cluster pairs | C0 | C1 | C2 |
| --- | --- | --- | --- |
| C0 | NA | 0.622 | 0.783 |
| C1 | 0.622 | NA | 1.0 |
| C2 | 0.783 | 1.0 | NA |

**TABLE 7.**
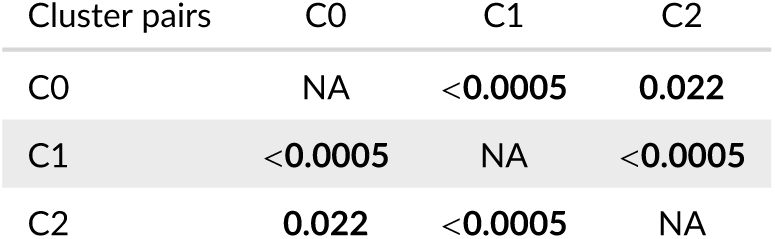
The significance for the schizophrenia stratum

### A.3 ICA-sFNC

**TABLE 8.**
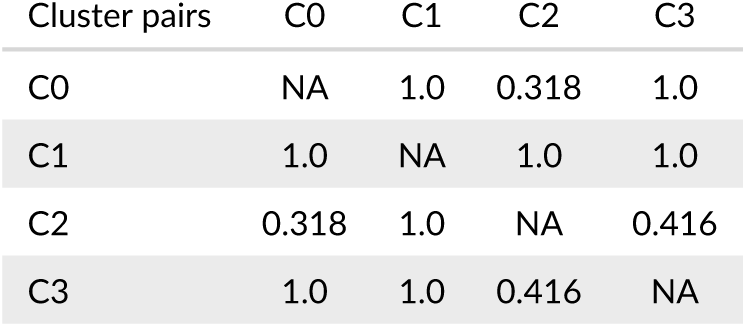
The significance for the sex stratum

| Cluster pairs | C0 | C1 | C2 | C3 |
| --- | --- | --- | --- | --- |
| C0 | NA | 1.0 | 0.318 | 1.0 |
| C1 | 1.0 | NA | 1.0 | 1.0 |
| C2 | 0.318 | 1.0 | NA | 0.416 |
| C3 | 1.0 | 1.0 | 0.416 | NA |

**TABLE 9.** The significance for the schizophrenia stratum

| Cluster pairs | C0 | C1 | C2 | C3 |
| --- | --- | --- | --- | --- |
| C0 | NA | <0.0005 | 0.084 | <0.0005 |
| C1 | <0.0005 | NA | <0.0005 | 1.0 |
| C2 | 0.084 | <0.0005 | NA | <0.0005 |
| C3 | <0.0005 | 1.0 | <0.0005 | NA |

### A.4 FA-ICA

**TABLE 10.**
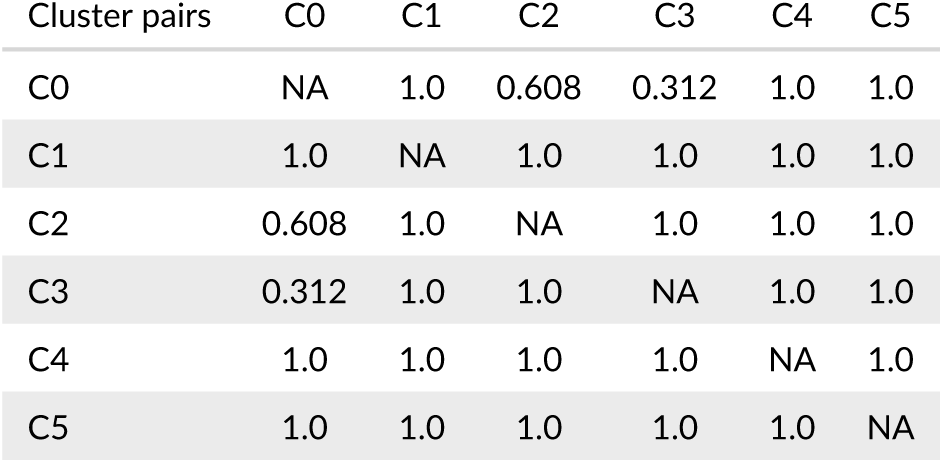
The significance for the sex stratum

| Cluster pairs | C0 | C1 | C2 | C3 | C4 | C5 |
| --- | --- | --- | --- | --- | --- | --- |
| C0 | NA | 1.0 | 0.608 | 0.312 | 1.0 | 1.0 |
| C1 | 1.0 | NA | 1.0 | 1.0 | 1.0 | 1.0 |
| C2 | 0.608 | 1.0 | NA | 1.0 | 1.0 | 1.0 |
| C3 | 0.312 | 1.0 | 1.0 | NA | 1.0 | 1.0 |
| C4 | 1.0 | 1.0 | 1.0 | 1.0 | NA | 1.0 |
| C5 | 1.0 | 1.0 | 1.0 | 1.0 | 1.0 | NA |

**TABLE 11.** The significance for the schizophrenia stratum

| Cluster pairs | C0 | C1 | C2 | C3 | C4 | C5 |
| --- | --- | --- | --- | --- | --- | --- |
| C0 | NA | **0.001** | 1.0 | 0.965 | 0.228 | 1.0 |
| C1 | **0.001** | NA | **<0.0005** | **0.037** | 1.0 | **0.003** |
| C2 | 1.0 | **<0.0005** | NA | 0.195 | 0.054 | 0.817 |
| C3 | 0.965 | **0.037** | 0.195 | NA | 0.935 | 1.0 |
| C4 | 0.228 | 1.0 | 0.054 | 0.935 | NA | 0.376 |
| C5 | 1.0 | **0.003** | 0.817 | 1.0 | 0.376 | NA |

### A.5 sMRI-FA

**TABLE 12.** The significance for the sex stratum

|  |  |  |  |  |
| --- | --- | --- | --- | --- |
| C0 | NA | **0.002** | 0.265 | **<0.0005** |
| C1 | **0.002** | NA | 0.265 | 1.0 |
| C2 | 0.265 | 0.265 | NA | **0.049** |
| C3 | **<0.0005** | 1.0 | **0.049** | NA |

**TABLE 13.** The significance for the schizophrenia stratum

| Cluster pairs | C0 | C1 | C2 | C3 |
| --- | --- | --- | --- | --- |
| C0 | NA | **<0.0005** | **<0.0005** | **0.002** |
| C1 | **<0.0005** | NA | 0.299 | **0.042** |
| C2 | **<0.0005** | 0.299 | NA | 1.0 |
| C3 | **0.002** | **0.042** | 1.0 | NA |

### A.6 sMRI-ICA

**TABLE 14.** The significance for the sex stratum

| Cluster pairs | C0 | C1 | C2 | C3 | C4 | C5 |
| --- | --- | --- | --- | --- | --- | --- |
| C0 | NA | 1.0 | 0.75 | 1.0 | 1.0 | 1.0 |
| C1 | 1.0 | NA | 1.0 | 0.279 | 1.0 | 0.559 |
| C2 | 0.75 | 1.0 | NA | 0.091 | 1.0 | 0.173 |
| C3 | 1.0 | 0.279 | 0.091 | NA | 0.174 | 1.0 |
| C4 | 1.0 | 1.0 | 1.0 | 0.174 | NA | 0.318 |
| C5 | 1.0 | 0.559 | 0.173 | 1.0 | 0.318 | NA |

**TABLE 15.** The significance for the schizophrenia stratum

| Cluster pairs | C0 | C1 | C2 | C3 | C4 | C5 |
| --- | --- | --- | --- | --- | --- | --- |
| C0 | NA | 1.0 | 0.059 | 0.077 | 0.358 | 1.0 |
| C1 | 1.0 | NA | **0.005** | 0.154 | 0.061 | 1.0 |
| C2 | 0.059 | **0.005** | NA | **<0.0005** | 1.0 | **0.005** |
| C3 | 0.077 | 0.154 | **<0.0005** | NA | **<0.0005** | 0.182 |
| C4 | 0.358 | 0.061 | 1.0 | **<0.0005** | NA | 0.06 |
| C5 | 1.0 | 1.0 | **0.005** | 0.182 | 0.06 | NA |

## B CHOOSING THE NUMBER OF MCPS FOR EACH MODALITY PAIR

To determine how many MCPs we select for each modality pair in our work, we use the elbow criterion. We look at the inertia averaged across 10 folds, see Figure 8. For each of the modality pairs, the number of MCPs seem to be between 3 and 6 MCPs, and four is the most common number of MCPs across modality pairs.

**FIGURE 8.**
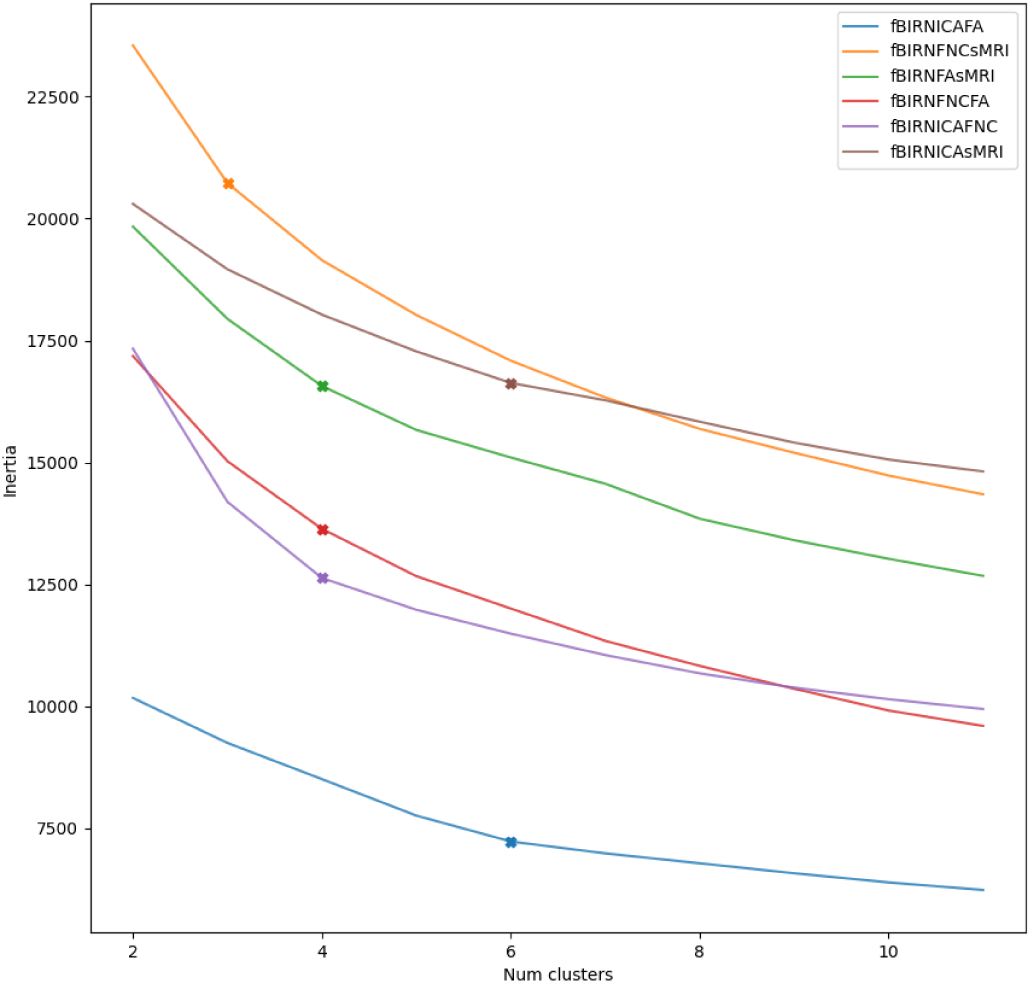
The inertia value vs the number of MCPs for each modality pair. For each of the modality pairs, we also visualize how many MCPs we use based on this curve with an x.

## C ROBUSTNESS AND CORRELATIONS TO SCHIZOPHRENIA FOR SHARED LATENT DIMENSIONS

To assess the robustness of shared latent dimensions across the 10 training folds, we calculated the correlations between latent dimensions across folds and then used the Hungarian algorithm [28, 29] to map all latent dimensions to the latent dimensions in fold 0. After calculating the best mapping, we averaged the correlations across folds, and only keep the latent dimensions with a correlation equal to or above 0.7, which we consider to be robust shared latent dimensions. For these robust latent dimensions, we calculate their correlation to schizophrenia, and we visualize the results in a heatmap in Figure 9. Note that quite a few of the shared latent dimensions for each modality pair are robust across folds. Furthermore, the correlation to schizophrenia for some of them is above 0.4. Note that the directionality of the correlation is arbitrary, and the most important aspect is the absolute value of the correlation. This accentuates the importance of training a model to find an explicitly shared space between modality pairs because they form a natural basis to analyze mental disorders like schizophrenia. In Section 3.5, we highlight two latent dimensions, namely FA-sFNC latent dimension 7, and sMRI-sFNC latent dimension 2 because of their interesting interpolation pattern from one end to the other.

**FIGURE 9.**
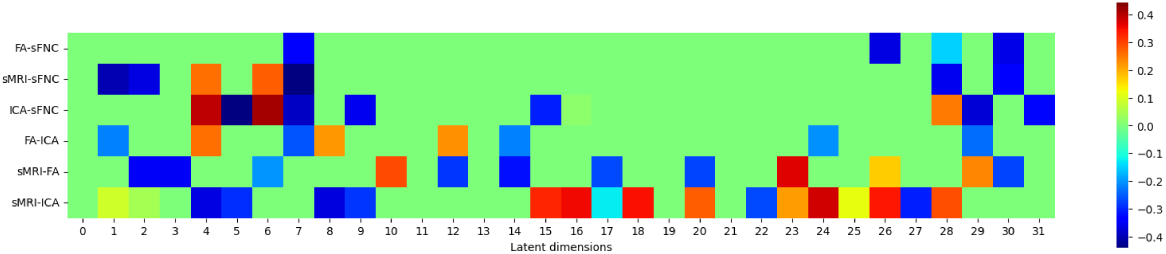
The correlation for robust shared latent dimensions to schizophrenia. For each of the modality pairs, we keep latent dimensions that are robust across folds (a correlation equal to or above 0.7), and calculate their correlation to schizophrenia.

